# Delayed Cord Clamping in China: A Cross-Sectional survey of Clinical Practices and Perceptions in Jiangsu Province

**DOI:** 10.1101/2025.05.18.24319380

**Authors:** Ling Yang, Yan Zhou, Lu Mei, Ning Gu, Hang Zhou, Wenwen Wang, Yimin Dai

## Abstract

**Objective:** To understand the awareness and practice of delayed cord clamping (DCC) among obstetric medical staff in Jiangsu Province.

**Design:** Cross-sectional study.

**Setting:** Obstetricians and midwives practicing in Jiangsu Province, China.

**Participants:** Create an online survey using the Wenjuanxing APP, and have the QR code link distributed by hospital administrators to the WeChat groups of obstetricians and midwives in hospitals, inviting them to participate in the survey.

**Outcome:** The questionnaire includes 28 items covering the respondents’ basic information, their knowledge and understanding of DCC, and its implementation status. Data analysis was based on frequencies and descriptive statistics.

**Results:** A total of 1001 people completed and submitted the questionnaire, with 866 included in the detailed analysis. This included 460 obstetricians (52.0%) and 406 midwives (48.1%). Among them, 50.7% reported being very familiar with DCC. 81.2% of respondents reported routinely implementing DCC, with midwives (91.4%) having a higher implementation rate than obstetricians (72.2%). In cases of cesarean section (89.8%), preterm birth (82.9%), and twin or multiple births (61.5%), most respondents chose to implement DCC. In cases of maternal infectious diseases, fewer chose to implement DCC: hepatitis B (31.9%), syphilis (24.4%), and HIV (18.4%). The main reasons for not implementing DCC were maternal bleeding or other emergencies (92.7%) and the need for neonatal resuscitation (88.8%).

**Conclusion:** The awareness and implementation rate of DCC among obstetric medical staff in Jiangsu Province are relatively high, but there are differences in specific details of cognition and practice, and no standard protocols have been formed. The application of DCC should continue to be promoted, and the early introduction of localized practice guidelines and standards should be advocated.

**Strengths and limitations of the study:**

☞ The survey covers all cities in Jiangsu Province, China, involving hospitals of different levels and natures. The work experience of the medical staff varies widely, which gives this survey a certain degree of representativeness.
☞ WeChat is the most widely used social software among Chinese people. Directly sending the survey to the medical staff WeChat groups in various hospitals can improve the response rate.
☞ Compared to other studies, this study’s survey items are more detailed, including the implementation of DCC in many special situations such as twins, cesarean sections, and maternal infectious diseases, providing a reference for practitioners.
☞ Jiangsu Province is located in the eastern coastal region of China and is one of the most economically developed regions in China. Medical staff there are relatively innovative in acquiring professional knowledge and more easily integrate it into practice, so the implementation rate of DCC should be higher than the national level.

## INTRODUCTION

Delayed cord clamping (DCC) is beneficial for the physiological transition from the fetal period to the neonatal period. It has been proven to reduce the transfusion rate, mortality and adverse neonatal outcomes of preterm infants, and improve the long-term iron reserves of term infants.^1-4^ Professional organizations WHO, ACOG and AAP have all issued recommendations for the implementation of DCC after the birth of preterm and term infants.^5-7^ The recently updated neonatal resuscitation guidelines of AAP also emphasized the cord management methods of neonates. The guidelines suggest that for term and preterm infants who do not require resuscitation, DCC (≥30s) is more beneficial than immediate cord clamping (ICC).^7^ Moreover, clinical research related to DCC is constantly updated, promoting the cognition and practice of this technology among medical staff. However, there are regional and hospital-specific differences in the practice of delayed cord clamping, influenced by various factors such as medical philosophy, clinical experience, and the specific conditions of the mother and newborn.

Current guidelines lack specific recommendations on the details of DCC implementation, including the optimal duration of DCC, the timing of oxytocin administration during DCC, and the placement of the newborn. Additionally, there are no clear recommendations for implementing DCC in special circumstances, such as in very preterm infants or in cases where the mother has complications or infectious diseases, which remains controversial.

Recognizing that research on DCC is still insufficient and that China lacks practical guidelines specifically on neonatal cord management methods, although some clinical guidelines recommend and advocate for DCC. We plan to conduct a cross-sectional survey. This study aims to understand the specific practices and experiences of DCC implementation in various delivery hospitals in Jiangsu Province, including the knowledge, attitudes, and practices of healthcare professionals regarding DCC.

## METHODS

### Study Design and Questionnaire Development

This study is a cross-sectional survey that has been approved by the Ethics Committee of Nanjing Drum Tower Hospital. The survey questionnaire was finalized by hospital management experts and senior obstetrics and neonatology specialists after multiple discussions and revisions. The questionnaire includes 28 items covering the respondents’ basic information, their knowledge and understanding of DCC, and its implementation status. The survey was conducted anonymously, with all questions being mandatory, and each device could only submit the questionnaire once. To assess the internal consistency of the questionnaire, SPSS software (version 25.0) was used to analyze part of the early collected data, yielding a Cronbach’s α coefficient of 0.939, indicating good internal consistency.

### Study Population and Data Collection

The study population consisted of obstetricians and midwives from more than 200 hospitals in Jiangsu Province. The questionnaire was created as an online survey using the Wenjuanxing APP, and the QR code link to the questionnaire was distributed to the WeChat groups of obstetricians and midwives by the hospital administrators. Participation was voluntary for all respondents. The questionnaire included standardized instructions, and designated personnel were responsible for the distribution and collection of the questionnaires to ensure the completeness and accuracy of the data. In further data analysis, people who were completely unaware of DCC, who had never implemented DCC and who had not been involved in delivery in the last three months were excluded.

### Sample Size Calculation

To ensure sufficient statistical power for the study, we performed a sample size calculation. Based on the average DCC implementation rate of 41.6% in 16 provinces of China as reported in the literature,^8^ with a significance level of 0.05, a statistical power of 0.80, and an allowable error of 0.05, the required sample size was calculated to be approximately 762. Considering that Jiangsu Province is one of the most developed regions in China and the actual implementation rate might be higher, we increased the sample size by 10%, resulting in a final sample size of 838.

### Statistical Analysis

After the survey was completed, the data were exported from the Wenjuanxing APP and analyzed using SPSS software(version 25.0). Continuous data with normal distribution were expressed as mean ± standard deviation and compared using the he independent samples Student’s t-test. Count data were summarized as numbers and percentages (%) and compared between the two groups with the Chi-squared or Fisher’s exact test. The Chi-squared test was utilized when the expected counts in any cell of a 2×2 table were greater than 5, while Fisher’s exact test was employed when the expected counts were less than 5. A difference was considered statistically significantly different at *p* < 0.05.

## RESULTS

A total of 1,001 respondents completed and submitted the questionnaire, among whom 22 indicated that they were completely unaware with DCC, resulting in a DCC awareness rate of 97.8%. There were 31 who never implemented DCC, leading to an implementation rate of 96.9%. Additionally, excluding 97 who had not participated in delivery work in the past three months, 866 respondents were included in the detailed analysis (There are duplications of these three types of people).

Consisting of 460 obstetricians (53.1%) and 406 midwives (46.9%) in 866 respondents. The majority were female (98.0%), from tertiary hospitals (83.3%), and general hospitals (74.7%), with most having 11-20 years of work experience (34.5%). The characteristics of the population are shown in Table 1.

**Table 1.** Demographic and characteristics of respondents (n = 866)

| Characteristics | n | % |
| --- | --- | --- |
| Gender |  |  |
| Female | 849 | 98.0 |
| Male | 17 | 2.0 |
| Professional category |  |  |
| Obstetrician | 460 | 53.1 |
| Midwife | 406 | 46.9 |
| Years of service |  |  |
| <5 | 54 | 6.2 |
| 5-10 | 215 | 24.8 |
| 11-20 | 299 | 34.5 |
| 21-30 | 184 | 21.3 |
| >30 | 114 | 13.2 |
| Hospital level |  |  |
| Tertiary hospital | 721 | 83.3 |
| Secondary hospital | 136 | 15.7 |
| Other | 9 | 1.0 |
| Hospital type |  |  |
| General hospital | 647 | 74.7 |
| Maternal-infant hospital | 165 | 19.1 |
| Chinese medicine hospital | 54 | 6.2 |
| Annual number of deliveries |  |  |
| <1000 | 272 | 31.4 |
| 1000-3000 | 417 | 48.2 |
| 3001-5000 | 86 | 9.9 |
| >5000 | 91 | 10.5 |
| Region |  |  |
| First-tier | 200 | 23.1 |
| Second-tier | 282 | 32.6 |
| Third-tier | 384 | 44.3 |

Region: Classification according to GDP (gross domestic product).

Among the 866 respondents included in the analysis, 50.7% reported being very familiar with DCC, while 49.3% reported being somewhat familiar with DCC. The majority (94.6%) believed that DCC is beneficial for newborns, and 51.7% of respondents expressed a desire for their departments to establish routine practices. Additionally, 93.1% of respondents indicated a desire for detailed guidelines or expert consensus to be issued. There was no difference in knowledge between obstetricians and midwives (Supplementary Table 1). 81.2% of the respondents reported routinely implementing DCC, with a higher implementation rate among midwives (91.4%) compared to obstetricians (72.2%). The proportion of departments with paper-based protocols was higher among midwives (56.9% vs. 28.9%) (Supplementary Table 2).

A comparison of obstetricians’ practices in different delivery methods showed that the proportion of those choosing 30-60 seconds for DCC implementation was significantly higher in cesarean sections than in vaginal deliveries (43.3% vs. 25.0%), while the proportion of those waiting for the umbilical cord to stop pulsating was significantly lower in cesarean sections than in vaginal deliveries (16.3% vs. 37.8%). Regarding the placement of newborns, most were placed on the mother’s abdomen during cesarean sections (47.6%), while during vaginal deliveries, they were more often placed on the thigh (47.2%) (Table 2).

**Table 2.** Obstetricians’ practice of DCC in different modes of delivery

|  | Delivery mode (n = 460) |  | P-value |
| --- | --- | --- | --- |
|  | Vaginal delivery | Cesarean delivery |  |
| Timing of DCC (s)<sup>a</sup> |  |  |  |
| 30-60 | 115 (25.0) | 199 (43.3) | <0.001 |
| 60-120 | 146 (31.7) | 152 (33.0) | 0.673 |
| 120-180 | 22 (4.8) | 19 (4.1) | 0.632 |
| >180 | 3 (0.7) | 3 (0.7) | - |
| Umbilical cord stops pulsing | 174 (37.8) | 75 (16.3) | <0.001 |
| Neonatal placement during DCC<sup>b</sup> |  |  |  |
| Chest | 105 (22.8) | 59 (12.8) | <0.001 |
| Abdomen | 166 (36.1) | 219 (47.6) | <0.001 |
| Thighs | 217 (47.2) | 177 (38.5) | 0.008 |
| Other | 22 (4.8) | 36 (7.8) | 0.058 |

a: Some people did not participate in cesarean delivery and therefore total percentage is less than 100%.
b: Multiple answers could be selected and therefore total percentage is more than 100%.

Compared to doctors who did not chose DCC in cesarean sections, those who choose DCC had a higher proportion of work experience at 21 to 30 years (30.0% vs. 10.9%), and a higher proportion of DCC implementation rates of more than 90.0% in the past three months (44.7% vs. 13.0%) (Supplementary Table 3).

In cases of cesarean sections (89.8%), preterm births (82.9%), and twin or multiple births (61.5%), most people choose to implement DCC. In the presence of maternal infectious diseases, fewer people choose to implement DCC, hepatitis B (31.9%), syphilis (24.4%), and HIV (18.4%). In cases of maternal complications, especially placental abruption, only 13.2% of people choose to implement DCC (Figure 1).

**Figure 1.**
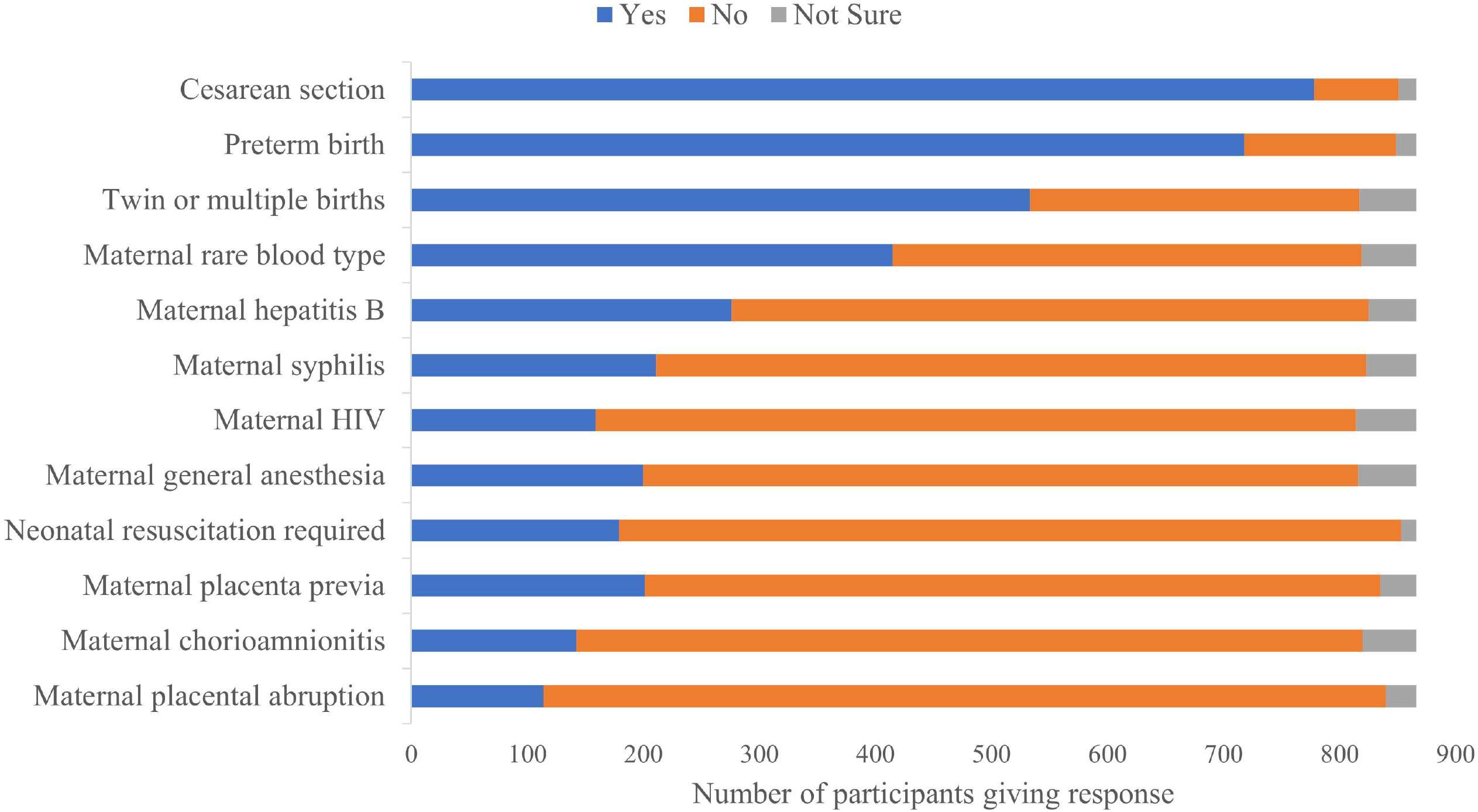
DCC implementation in special cases.

In hospitals where DCC is implemented, the main reasons for not implementing DCC are emergency situations such as maternal hemorrhage (92.7%) and the need for neonatal resuscitation (88.8%) (Figure 2). A survey was also conducted among individuals who are aware of DCC but do not implement it, revealing that the main reasons are the lack of routine protocols in the department (58.1%), insufficient manpower (38.7%), lack of knowledge about DCC (35.5%), and difficult in operation (35.5%) (Supplementary Table 4).

**Figure 2.**
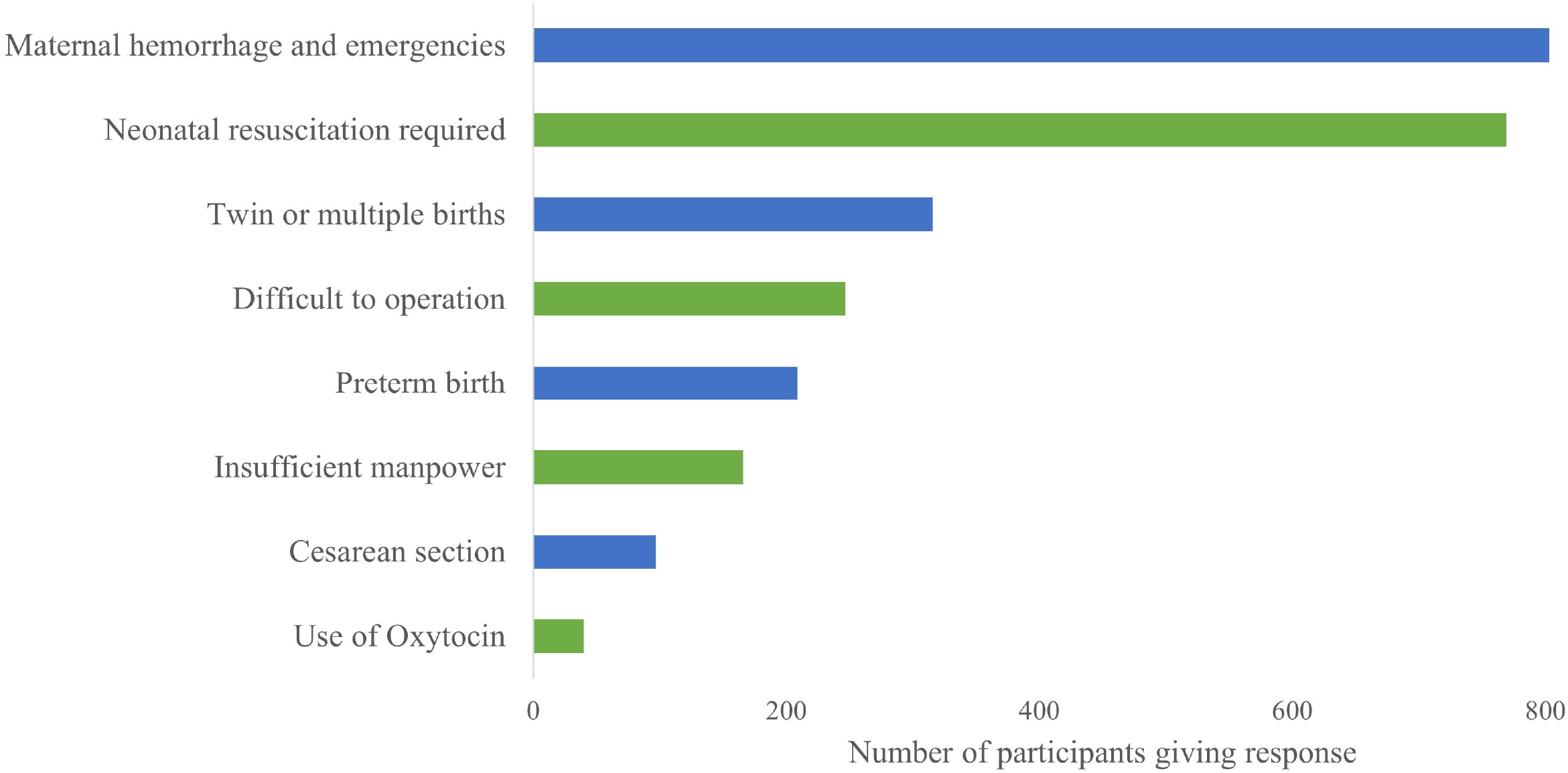
The main reasons for not implementing DCC.

## DISCUSSION

This study used anonymous questionnaires to collect information on the knowledge and practices of DCC among healthcare personnel in various maternity hospitals in Jiangsu Province. The results indicate that the awareness and implementation rates of DCC among obstetric healthcare personnel in Jiangsu are relatively high, though there are differences in practice details. Considering that the survey involved hospitals of different levels and natures, with a wide range of years of service among healthcare personnel, the findings should be fairly representative.

Our research shows that 81.2% of obstetric healthcare personnel routinely implement DCC in their delivery work, which is significantly higher than the DCC implementation rate (41.6%) found in a 2020 survey across 16 provinces in China,^8^ and close to a 2020 survey result in the United States (81.7%).^9^ The high implementation rate can be attributed to the fact that most participants in this study come from tertiary hospitals, with the majority having over 10 years of service. Additionally, Jiangsu is an economically developed region in China, where healthcare personnel have better access to innovative professional knowledge and are more likely to integrate it into practice.

The ACOG first recommended DCC in 2012 and updated it in 2017.^6,10^ An epidemiological survey conducted in the United States in 2018 found that the implementation rate of DCC had significantly increased compared to the 2014 survey results, highlighting the importance of formal guidelines for clinical practice.^11^ Although the DCC implementation rate in this study is high, few hospitals have established formal protocols, with only 42.0% having written guidelines. Moreover, our country lacks professional practice guidance, and the absence of departmental protocols is a primary reason for not implementing DCC. In clinical practice, midwives generally follow established protocols, while doctors pursue cutting-edge knowledge. Our study indeed found that departments with written guidelines had higher routine implementation rates, and most doctors’ awareness of DCC practices likely stems from international guidelines and peer learning.

Despite multiple meta-analyses, the optimal duration for DCC remains undetermined. Most studies recommend a minimum duration of 60 seconds, with some extending up to 180 seconds. A small sample study indicated that DCC could last up to 4 minutes or longer when the newborn is stable, but many outcome indicators showed no significant differences compared to the 30-60 seconds group. Conversely, longer durations could lower umbilical blood pH, increase the risk of hypothermia (48.5%), and elevate the risk of hyperbilirubinemia requiring phototherapy (94.6%).^12,13^ A meta-analysis of RCTs showed that DCC beyond 60 seconds in full-term infants did not improve morbidity and mortality but increased the risk of hyperbilirubinemia requiring phototherapy.^14^ The latest meta-analysis, including 47 clinical trials, suggested that DCC for ≥ 120 seconds significantly reduced preterm mortality before discharge compared to ICC.^3^ China’s “ Normal Delivery Guidelines” recommend clamping the umbilical cord at least 60 seconds after birth or waiting until the umbilical cord pulsation stops for full-term infants not needing resuscitation.^15^ Our study shows that 43.2% of respondents (vaginal delivery) believe the cord should be clamped after the pulsation stops. The optimal duration for DCC remains unclear and requires more research evidence.

Although there is substantial evidence indicating the benefits of DCC for newborns, there is limited research on cesarean deliveries, especially concerning maternal outcomes. Obstetricians still worry that the waiting time for DCC might affect surgical procedures and increase maternal blood loss. We surveyed healthcare personnel’s attitudes towards DCC in cesarean sections, finding that 90.0% of obstetricians would choose DCC during cesarean sections. Those who did not choose DCC generally had shorter years of service, lower recent implementation rates, worked in lower-level hospitals, and had fewer annual deliveries (Supplementary Table 3). The duration of DCC chosen during cesarean sections was shorter than that in vaginal deliveries. An RCT comparing 60 seconds of DCC to ECC found no significant difference in maternal hemoglobin levels on the first postoperative day.^16^ Other literature has confirmed the safety of 90-120 seconds of DCC for cesarean section mothers.^17^ Regarding positioning, the 2022 SOGC recommends placing the newborn at the vaginal introitus or below the uterine incision level.^18^ A meta-analysis of preterm infants did not determine the optimal position for the newborn at delivery, but considering gravitational effects, placing the newborn lower seems reasonable.^2^ The ACOG suggests that newborns during cesarean sections can be placed on the mother’s abdomen or legs, or by the surgeon near the placental level until the cord is clamped.^6^ Overall, current DCC practices during cesarean sections are greatly influenced by doctors’ clinical experience, and more detailed and compelling evidence is needed to guide practice.

Regarding the practice survey in special situations, most respondents would choose DCC in cesarean and preterm cases, but many were confused about whether to implement DCC in cases of maternal infectious diseases. The WHO recommends DCC for all women, including HIV-positive mothers and those with unknown HIV status.^5^ Italy suggests DCC for newborns of mothers with adequately treated HIV and low viral loads during pregnancy for 60 seconds.^19^ A clinical study confirmed that newborns of HIV-positive mothers remained negative for the virus after DCC.^20^ In our survey, 75.6% of respondents chose not to perform DCC for HIV-positive mothers. Respondents also had a conservative attitude towards DCC for mothers with HBV and syphilis, although no relevant literature was found. More research evidence is needed for DCC in infectious disease cases.

Research on DCC in twin births is limited, and existing studies have not stratified different types of twins. Limited evidence suggests that DCC in twins reduces the need for neonatal transfusions, surfactant use, and the risks of intubation and mechanical ventilation.^2,21^ The 2022 SOGC recommends considering DCC for twins unless contraindicated.^18^ In our study, 61.5% of respondents chose to perform DCC for twins or multiples. Our team is currently conducting an RCT on DCC in twins, including monochorionic twins, which may provide evidence for developing localized practice guidelines.

The SOGC also suggests that for centers with appropriate experience and equipment, using the intact umbilical cord for longer stabilization or resuscitation of preterm and full-term infants is feasible, although larger trials are needed to understand the benefits and risks.^18^ Our study shows that 20.7% of respondents believe DCC can be performed when neonatal resuscitation is needed, but in practice, 77.8% chose not to perform DCC due to the need for resuscitation. Similarly, fewer personnel chose to perform DCC under maternal complications or general anesthesia. DCC is also recommended in the latest postpartum hemorrhage guidelines in China, however our survey shows that maternal haemorrhage is the most important reason for not having DCC.^22^ This is related to our medical conditions, as most institutions may not have the facilities for bedside resuscitation with an intact cord. In emergencies such as placenta previa or placental abruption, DCC implementation is hindered by operational difficulties or manpower shortages.

The limitations of this study include the inability to determine the number of individuals who read but did not submit the questionnaire after we published the QR code, making it impossible to calculate the response rate. There may be a selection bias, as those knowledgeable about DCC might be more willing to complete the survey, potentially leading to an overestimation of the actual implementation rate.

## CONCLUSION

Overall, the implementation rate of DCC among obstetric healthcare personnel in Jiangsu Province, China, is high, and their attitude towards DCC is positive. Even those who did not routinely implement DCC expressed willingness to do so in the future. DCC is a free and simple intervention with the potential to benefit millions of newborns born each year in China. However, more reliable research evidence and professional guidelines are needed to promote and guide our clinical practice. This survey may provide a basis for establishing routine clinical protocols in maternity hospitals and for professional organizations to issue guidelines on umbilical cord management for newborns.

## Authors and Contributions

Ling Yang and Lu Mei analyzed the data and wrote the manuscript. Hang Zhou, Yan Zhou and Ning Gu distributed the questionnaires and collected the data. Wenwen Wang and Yimin Dai conceived and designed the study. All authors contributed to editorial changes and approved the final version of the manuscript.

## Patient and public involvement

Patients and/or the public were not involved in the design, or conduct, or reporting, or dissemination plans of this research.

## Patient consent for publication

Not applicable.

## Ethical approval

This study was approved by Medical Ethics Committee of Tower Hospital Affiliated to Nanjing University Medical school (Ethics code: 2024-142-01). All participants were informed about the study’s aims and procedures, written consent was obtained from patients to participate in the study. The procedures used in this study adhere to the tenets of the Declaration of Helsinki.

## Provenance and peer review

Not commissioned; externally peer reviewed.

## Data availability statement

Data are available upon reasonable request. Interested parties can contact the corresponding author with their inquiry.

## Acknowledgements

We gratefully acknowledge the support from the participating hospitals and all the healthcare professionals who contributed their valuable time to this survey.

