## Supplementary Table for "Delayed Cord Clamping in China: A Cross-Sectional survey of Clinical Practices and Perceptions in Jiangsu Province"

| Supplementary Table 1. Respondents' knowledge of DCC (n = 866) | | | |
| --- | --- | --- | --- |
| Knowledge situation | Total  (n = 866) | Obstetricians  (n = 460) | Midwives  (n = 406) |
| Knowledge level |  |  |  |
| Highly understood | 439 (50.7) | 222 (48.3) | 217 (53.4) |
| Partial understood | 427 (49.3) | 238 (51.7) | 189 (46.6) |
| Views on DCC |  |  |  |
| Benefits for newborns | 828 (94.6) | 432 (93.9) | 396 (97.5) |
| Benefits not advantageous | 70 (8.6) | 31 (6.7) | 39 (9.6) |
| Pros and Cons, Difficult Choice | 102 (12.7) | 42 (9.1) | 60 (14.8) |
| No benefits for the mother | 43 (5.0) | 23 (5.0) | 20 (5.0) |
| Outlook for DCC |  |  |  |
| Need for detailed guidelines or expert consensus | 806 (93.1) | 433 (94.1) | 373 (91.9) |
| Need for establish routines in our department | 506 (51.7) | 239 (52.0) | 238 (58.6) |
| Need for more research to support DCC | 750 (76.6) | 344 (74.8) | 369 (90.9) |

| Supplementary Table 2. Practice of DCC among respondents and their hospitals (n = 866) | | | |
| --- | --- | --- | --- |
|  | Total  (n = 866) | Obstetricians (n = 460) | Midwives  (n = 406) |
| Implementation |  |  |  |
| Routine | 703 (81.2) | 332 (72.2) | 371 (91.4) |
| Non-routine | 163 (18.8) | 128 (27.8) | 35 (8.6) |
| Practice Standards |  |  |  |
| Paper-based protocols | 364 (42.0) | 133 (28.9) | 231 (56.9) |
| Orally imparted | 400 (46.2) | 263 (57.2) | 137 (33.7) |
| No standards | 102 (11.8) | 64 (13.9) | 38 (9.4) |
| Implementation rate in the last 3 months (%) |  |  |  |
| >90 | 454 (52.4) | 191 (41.5) | 263 (64.8) |
| 51-90 | 263 (30.4) | 155 (33.7) | 108 (26.6) |
| 20-50 | 80 (9.2) | 59 (12.8) | 21 (5.2) |
| <20 | 69 (8.0) | 55 (12.0) | 14 (3.4) |
| Implementation time of vaginal delivery DCC (s)^a^ |  |  |  |
| 30-60 | 161 (18.6) | 115 (25.0) | 46 (11.3) |
| 60-120 | 230 (26.6) | 138 (30.0) | 92 (22.7) |
| 120-180 | 36 (4.2) | 20 (4.4) | 16 (3.9) |
| >180 | 10 (1.2) | 3 (0.7) | 7 (1.7) |
| Umbilical cord stops pulsating | 374 (43.2) | 142 (30.9) | 232 (57.1) |
| Timing of Oxytocin in DCC |  |  |  |
| Before cord clamping | 373 (43.1) | 127 (27.6) | 246 (60.6) |
| After cord clamping | 358 (41.3) | 272 (59.1) | 86 (21.2) |
| Both | 135 (15.6) | 61 (13.3) | 74 (18.2) |
| Who watches newborns at DCC^b^ |  |  |  |
| Obstetricians | 407 | 272 (59.1) | 135(33.3) |
| Neonatologist | 139 | 98 (21.3) | 41 (10.1) |
| Nurses | 79 | 43 (9.4) | 36 (8.9) |
| Midwives | 764 | 367 (79.8) | 397 (97.8) |
| Neonatal care measures in the event of DCC |  |  |  |
| Dry and wrap with a pre-heated towel | 610 (70.4) | 263 (57.2) | 347 (85.5) |
| No drying, wrap with a pre-heated towel | 41 (4.7) | 27 (5.9) | 14 (3.5) |
| Dry, no wrapping | 187 (21.6) | 144 (31.3) | 43 (10.6) |
| No drying, no wrapping | 28 (3.2) | 26 (5.7) | 2 (0.5) |

a: Some people did not make a choice and therefore total percentage is less than 100%.

b: Multiple answers could be selected and therefore total percentage is more than 100%.

| Supplementary Table 3. Comparison of Obstetricians Characteristics in Choosing DCC during Cesarean Delivery | | | |
| --- | --- | --- | --- |
| Obstetricians' characteristics | DCC  (n = 414) | Non-DCC  (n = 46) | P-value |
| Years of service |  |  |  |
| <5 | 18 (4.3) | 4 (8.7) | 0.260 |
| 5-10 | 55 (13.3) | 11 (23.9) | 0.051 |
| 11-20 | 139 (33.6) | 10 (21.7) | 0.104 |
| 21-30 | 124 (30.0) | 5 (10.9) | 0.006 |
| >30 | 78 (18.8) | 10 (21.7) | 0.635 |
| Hospital level |  |  |  |
| Tertiary | 323 (78.0) | 33 (71.7) | 0.334 |
| Secondary | 85 (20.5) | 12 (26.1) | 0.381 |
| Other | 6 (1.4) | 1 (2.2) | 0.703 |
| Type of hospital |  |  |  |
| General hospitals | 306 (73.9) | 38 (82.6) | 0.198 |
| Maternal-infant hospital | 73 (17.6) | 8 (17.4) | 0.967 |
| Chinese medicine hospital | 35 (8.5) | 0 | - |
| Annual number of deliveries |  |  |  |
| <1000 | 147 (35.5) | 17 (37.0) | 0.846 |
| 1000-3000 | 188 (45.4) | 23 (50.0) | 0.553 |
| 3001-5000 | 42 (10.1) | 4 (8.7) | 0.746 |
| >5000 | 37 (8.9) | 2 (4.3) | 0.289 |
| Implementation rate in the last 3 months (%) |  |  |  |
| >90 | 185 (44.7) | 6 (13.0) | <0.001 |
| 51-90 | 148 (35.7) | 7 (15.2) | 0.005 |
| 20-50 | 46 (11.1) | 13 (28.3) | 0.001 |
| <20 | 35 (8.5) | 20 (43.5) | <0.001 |

| Supplementary Table 4. Reasons for not implementing DCC | | |
| --- | --- | --- |
| Reasons for not implementing DCC (n = 31) ^a^ | n | % |
| No routine in the department | 18 | 58.1 |
| Not considered good for newborn | 1 | 3.2 |
| Not considered good for mother | 1 | 3.2 |
| Don't understand DCC | 11 | 35.5 |
| Difficult to operation | 11 | 35.5 |
| Insufficient manpower | 12 | 38.7 |

a: Multiple answers could be selected and therefore total percentage is more than 100%.
